# Ethnic inequities in multimorbidity among people with psychosis: A retrospective cohort study

**DOI:** 10.1101/2022.02.16.22271050

**Authors:** Daniela Fonseca de Freitas, Megan Pritchard, Hitesh Shetty, Mizanur Khondoker, James Nazroo, Richard D. Hayes, Kamaldeep Bhui

## Abstract

**Background:** Research shows persistent ethnic inequities in mental health experiences and outcomes, with a higher incidence of illnesses among minoritised ethnic groups. People with psychosis have a higher risk of multiple long-term conditions (MLTC; multimorbidity). However, there is limited research regarding ethnic inequalities in multimorbidity in people with a schizophrenia spectrum disorder. This study investigates ethnic disparities in physical health multimorbidity in a cohort of people with psychosis.

**Methods:** In this retrospective cohort study, using the Clinical Records Interactive Search (CRIS) system, we identified service-users of the South London and Maudsley NHS Trust with a schizophrenia spectrum disorder, and then additional diagnoses of diabetes, hypertension, low blood pressure, overweight or obesity, and rheumatoid arthritis. Multinomial logistic regression was then used to investigate ethnic inequities in odds of multimorbidity (psychosis plus one physical health condition), as well as multimorbidity severity (having one or two physical health conditions, or three or more conditions), compared with no additional health conditions (no multimorbidity). The regression models adjusted for age and duration of care and investigated the influence of gender and area-level deprivation.

**Results:** On a sample of 20,800 service-users with psychosis, aged 13-65, ethnic differences were observed in the odds for multimorbidity. Compared to White British people, higher odds of multimorbidity were found for people of Black African [aOR=1.41, 95%CI (1.23 - 1.56)], Black Caribbean [aOR=1.79, 95%CI (1.58 – 2.03)], and Black British [aOR=1.64, 95%CI (1.49 - 1.81)] ethnicity. Reduced odds were observed among people of Chinese [aOR=0.61, 95%CI (0.43 – 0.88)] and Other ethnicities [aOR=0.67, 95%CI (0.59 – 0.76)]. Increased odds for severe multimorbidity (three or more physical health conditions) were also observed for people of any Black background.

**Conclusions:** Ethnic inequities are observed for multimorbidity among people with psychosis. Further research is needed to understand the aetiology and impact of these inequities. These findings support the provision of integrated health care interventions and public health preventive policies and actions.

## INTRODUCTION

Ethnic inequities^1^ in health have been reported (El-Sayed et al., 2011; J. Y. Nazroo et al., 2009; Sproston & Mindell, 2006; Watkinson et al., 2021). For example, previous studies suggest a higher prevalence of obesity, hypertension and cardiovascular disease among Black people in the UK (El-Sayed et al., 2011; J. Y. Nazroo et al., 2009; Schofield et al., 2011; Sproston & Mindell, 2006); higher risk of diabetes has been observed among people of Black Caribbean, Indian, Pakistani, Bangladeshi and Chinese background (J. Y. Nazroo et al., 2009). The prevalence of multiple long-term conditions (MLTC, or multimorbidity) was observed to be higher in almost all minoritised ethnic groups in the UK (Bisquera et al., 2021; Watkinson et al., 2021). Also, health related quality of life and self-rated health, a predictor of mortality, is poorer in several minoritised ethnic groups (Evandrou et al., 2016; Watkinson et al., 2021). Ethnic health inequities have been largely associated with differences in socioeconomic status (income, education, occupation) (Dubath et al., 2021; James Y. Nazroo & Williams, 2006). However, evidence suggests that ethnic inequities persist after controlling for socioeconomic factors, though such models may contain residual confounding (Evandrou et al., 2016; Larsen et al., 2017; James Y. Nazroo, 1998; James Y. Nazroo & Williams, 2006; Verest et al., 2019; Watkinson et al., 2021).

People with severe mental illness (SMI, including psychosis and bipolar disorder) have an elevated risk of multiple long-term conditions (or multimorbidity), with higher incidence of diabetes, hypertension, asthma, chronic obstructive pulmonary disease and chronic kidney disease (Rodrigues et al., 2021; Stubbs et al., 2016; Woodhead et al., 2014). However, there is limited evidence on ethnic inequities in multimorbidity among people with psychosis. A recent study reported that people from a minoritised ethnic background show an increased risk of diabetes in the first year of psychotic illness, while no major changes were observed in White people (Gaughran et al., 2019). No ethnicity-related difference in overall weight (body mass index), or changes in it, were observed. However, increases in central obesity (mean waist circumference) were observed among minoritised ethnic women and White men, but no major changes were observed in minoritised ethnic men or White women (Gaughran et al., 2019).

This study aims to investigate ethnic inequities in multimorbidity, and multimorbidity severity, among a cohort of people with psychosis. Acknowledging differences in multimorbidity related to gender (Gaughran et al., 2019; Head et al., 2021; Larsen et al., 2017; Watkinson et al., 2021) and socioeconomic deprivation (Dubath et al., 2021; Head et al., 2021; Marmot et al., 2020), we investigate interaction effects. We hypothesise that people of a minoritised ethnic background present higher multimorbidity than White British people and that this is amplified for minoritised ethnic women and minoritised ethnic people living in more deprived areas.

## METHOD

### Setting

This retrospective cohort study used data from the electronic health records (EHRs) of the South London and Maudsley (SLaM) National Health Service (NHS) Foundation Trust. SLaM’s catchment area comprises four ethnically diverse London boroughs (Southwark, Lewisham, Lambeth, and Croydon) covering 1.3 million people. Access to clinical records was obtained via the Clinical Record Interactive Search (CRIS) system (Perera et al., 2016; Stewart et al., 2009). CRIS was established under a robust data protection and governance framework and received approval from the Oxford C Research Ethics Committee (18/SC/0372) to be used as a de-identified dataset for secondary data analysis (Perera et al., 2016; Stewart et al., 2009). Projects using CRIS are submitted for approval to a service-user led oversight committee; this project’s reference is 20-075.

At the time of writing, CRIS enabled access to the de-identified information, in the free-text and structured fields, of the clinical records of over 400,000 service-users. One way to efficiently access and code information on the free-text fields is by using Natural Language Processing (NLP) algorithms, which eliminate the need for researchers to read and code large volumes of text (CRIS NLP Service, 2021; Jackson et al., 2017; Perera et al., 2016). These algorithms analyse text and identify the condition of interest, distinguishing true events from false positives (e.g., notes about a condition without the service-user having that condition) (Jackson et al., 2017; Perera et al., 2016). More information on the NLP algorithms used with CRIS can be found on CRIS NLP Service webpage (CRIS NLP Service, 2021). The study is reported according the RECORD statement (Benchimol et al., 2015).

### Participants

SLaM service-users who meet the following inclusion criteria: (i) had a schizophrenia spectrum disorder (WHO ICD-10 codes F20–F29, on structured and free-text fields of the electronic records (CRIS NLP Service, 2021)), diagnosed between 1 January 2007 and 31 December 2020, (ii) were aged between 13 and 65 years at the time of that diagnosis, (iii) had at least one clinical event between 1 January 2007 and 31 December 2020, and (iv) had information regarding ethnic identity.

### Variables

#### Multimorbidity and multimorbidity severity

Based on the current information available on CRIS we were able to identify the prevalence of six LTCs: asthma, bronchitis, diabetes, hypertension, low blood pressure, overweight or obesity, and rheumatoid arthritis. Multimorbidity is commonly defined as the coexistence of two or more long-term health conditions (Johnston et al., 2019; NICE (National Institute for Health and Care Excellence), 2016); so among the cohort of people with a schizophrenia spectrum disorder, multimorbidity was recognised when there was evidence of at least one physical health condition. To assess severity of multimorbidity, we further analysed the odds for the presence of one or two physical health conditions, and presence of three or more physical health conditions.

The information regarding physical health conditions relied on the use of NLP algorithms (CRIS NLP Service, 2021) and this information was collected during the study’s observation period, from 01/01/2007 to 31/12/2020. All algorithms used in this study showed good performance, with precision over 90% and recall over 76% (except for bronchitis, where recall was observed to be 48%) (CRIS NLP Service, 2021). The NLP algorithm identifying asthma analyses mentions of asthma in text. Similarly, bronchitis identification is based on statements of chronic obstructive pulmonary disease, chronic bronchitis, centrilobular emphysema or other terms related to bronchitis. Information on diabetes was derived from a combination of four sources of information: physical health forms, results from blood analysis where the haemoglobin A1c (HbA_1c_) test (which measures average blood sugar levels) indicated diabetes, and NPL algorithms to identify medications for diabetes or in-text evidence of diabetes. Hypertension was identified via the use of two NLP algorithms: one that focuses on mentions of hypertension problems in the text, and another that analyses information regarding blood pressure; high blood pressure was identified when the systolic pressure was above 140mmHg, or the diastolic pressure was above 90mmHg. Low blood pressure was determined when systolic pressure was below 90 mmHg or the diastolic pressure was below 60 mmHg. To identify overweight or obesity, at any point during the observation period, we used an NLP algorithm that identified Body Mass Index (BMI) values; the information was coded as evidence of overweight or obesity when the BMI was between 25 and 50. Data regarding rheumatoid arthritis was derived from a NLP algorithm identifying mentions of rheumatoid arthritis in text (CRIS NLP Service, 2021).

### Exposure and control variables

Ethnicity was derived from 16 NHS categories. White British ethnicity was used as a reference category in the analyses. Because of small sample sizes, we have grouped mixed ethnic identities (e.g., Black African and White ethnicity). Other sociodemographic data comprised gender, age at diagnosis, and area-level deprivation. Area-level deprivation was based on the English Indices of Deprivation 2015 (Department for Communities and Local Government, 2015); we retrieved the decile of the index of multiple deprivation attributed to the lower-layer super output area of the service-users address at the diagnosis date. Based on these deciles, and considering the high deprivation level in the SLaM catchment area, four groups were derived: most deprived (comprising people living in deciles 1 and 2, corresponding the 20% most deprived areas in England), second most deprived (comprising people living in decile 3 and 4), middle deprivation (comprising people living in decile 5 and 6), least deprived (comprising deciles 7 to 10, corresponding to the 40% least deprived areas). To control for the surveillance bias associated with the availability of information, we included a measure that estimated each individual’s observation period, commencing with the date of the first diagnosis of SSD after 2007 and finishing at the end of the study’s observation period (31/12/2020), or date of death.

### Statistical analysis

Logistic regression analyses were conducted to investigate the association between ethnicity and multimorbidity (one or more physical health conditions) while controlling for other sociodemographic information and the observation period’s length. Multinomial regression analyses were used to investigate association between ethnicity and multimorbidity morbidity severity, considering no multimorbidity, 1 or 2 physical health conditions, and 3 or more physical health conditions. We tested interactions between ethnicity and gender and ethnicity and area-level deprivation in the predictive model for multimorbidity by using the likelihood ratio test comparing the models with and without the interaction terms, with adjustments for age, area-level deprivation and gender, respectively. Statistical analyses were conducted using STATA 15 (StataCorp, 2017).

## RESULTS

### Participants

The sample comprises 20,800 service-users who met the inclusion criteria; 10% of those meeting the inclusion criteria were excluded because of missing or uncertain ethnicity data. The largest ethnic group was people of White British ethnicity (35%), followed by Black British or other Black background (15%), Black African (14%), Black Caribbean (9%) and Other White ethnicity (9%) (Table 1). The majority of the cohort comprised men (61%), and the mean age at diagnosis was 35.5 years. Most people lived in highly deprived neighbourhoods (38% living in the 20% most deprived areas in England). Further information on the sample description is presented in Table 1.

**Table 1.**
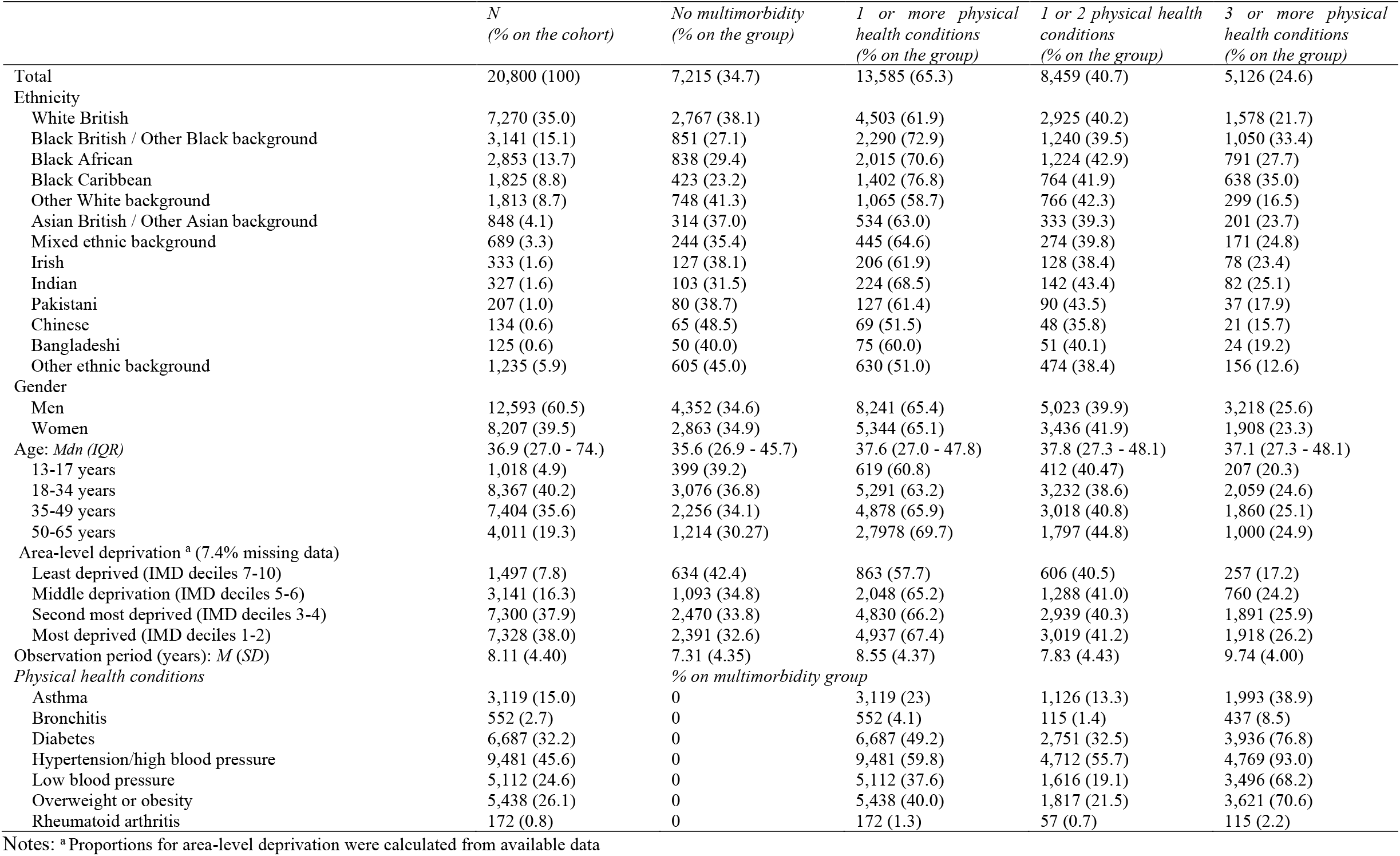
Characteristics of the study’s cohort

|  | *N*<br>(% on the cohort) | *No multimorbidity*<br>(% on the group) | *1 or more physical health conditions*<br>(% on the group) | *1 or 2 physical health conditions*<br>(% on the group) | *3 or more physical health conditions*<br>(% on the group) |
| --- | --- | --- | --- | --- | --- |
| Total | 20,800 (100) | 7,215 (34.7) | 13,585 (65.3) | 8,459 (40.7) | 5,126 (24.6) |
| Ethnicity |  |  |  |  |  |
| White British | 7,270 (35.0) | 2,767 (38.1) | 4,503 (61.9) | 2,925 (40.2) | 1,578 (21.7) |
| Black British / Other Black background | 3,141 (15.1) | 851 (27.1) | 2,290 (72.9) | 1,240 (39.5) | 1,050 (33.4) |
| Black African | 2,853 (13.7) | 838 (29.4) | 2,015 (70.6) | 1,224 (42.9) | 791 (27.7) |
| Black Caribbean | 1,825 (8.8) | 423 (23.2) | 1,402 (76.8) | 764 (41.9) | 638 (35.0) |
| Other White background | 1,813 (8.7) | 748 (41.3) | 1,065 (58.7) | 766 (42.3) | 299 (16.5) |
| Asian British / Other Asian background | 848 (4.1) | 314 (37.0) | 534 (63.0) | 333 (39.3) | 201 (23.7) |
| Mixed ethnic background | 689 (3.3) | 244 (35.4) | 445 (64.6) | 274 (39.8) | 171 (24.8) |
| Irish | 333 (1.6) | 127 (38.1) | 206 (61.9) | 128 (38.4) | 78 (23.4) |
| Indian | 327 (1.6) | 103 (31.5) | 224 (68.5) | 142 (43.4) | 82 (25.1) |
| Pakistani | 207 (1.0) | 80 (38.7) | 127 (61.4) | 90 (43.5) | 37 (17.9) |
| Chinese | 134 (0.6) | 65 (48.5) | 69 (51.5) | 48 (35.8) | 21 (15.7) |
| Bangladeshi | 125 (0.6) | 50 (40.0) | 75 (60.0) | 51 (40.1) | 24 (19.2) |
| Other ethnic background | 1,235 (5.9) | 605 (45.0) | 630 (51.0) | 474 (38.4) | 156 (12.6) |
| Gender |  |  |  |  |  |
| Men | 12,593 (60.5) | 4,352 (34.6) | 8,241 (65.4) | 5,023 (39.9) | 3,218 (25.6) |
| Women | 8,207 (39.5) | 2,863 (34.9) | 5,344 (65.1) | 3,436 (41.9) | 1,908 (23.3) |
| Age: *Mdn (IQR)* | 36.9 (27.0 - 74.) | 35.6 (26.9 - 45.7) | 37.6 (27.0 - 47.8) | 37.8 (27.3 - 48.1) | 37.1 (27.3 - 48.1) |
| 13-17 years | 1,018 (4.9) | 399 (39.2) | 619 (60.8) | 412 (40.47) | 207 (20.3) |
| 18-34 years | 8,367 (40.2) | 3,076 (36.8) | 5,291 (63.2) | 3,232 (38.6) | 2,059 (24.6) |
| 35-49 years | 7,404 (35.6) | 2,256 (34.1) | 4,878 (65.9) | 3,018 (40.8) | 1,860 (25.1) |
| 50-65 years | 4,011 (19.3) | 1,214 (30.27) | 2,797 (69.7) | 1,797 (44.8) | 1,000 (24.9) |
| Area-level deprivation <sup>a</sup> (7.4% missing data) |  |  |  |  |  |
| Least deprived (IMD deciles 7-10) | 1,497 (7.8) | 634 (42.4) | 863 (57.7) | 606 (40.5) | 257 (17.2) |
| Middle deprivation (IMD deciles 5-6) | 3,141 (16.3) | 1,093 (34.8) | 2,048 (65.2) | 1,288 (41.0) | 760 (24.2) |
| Second most deprived (IMD deciles 3-4) | 7,300 (37.9) | 2,470 (33.8) | 4,830 (66.2) | 2,939 (40.3) | 1,891 (25.9) |
| Most deprived (IMD deciles 1-2) | 7,328 (38.0) | 2,391 (32.6) | 4,937 (67.4) | 3,019 (41.2) | 1,918 (26.2) |
| Observation period (years): *M (SD)* | 8.11 (4.40) | 7.31 (4.35) | 8.55 (4.37) | 7.83 (4.43) | 9.74 (4.00) |
| *Physical health conditions* |  | *% on multimorbidity group* |  |  |  |
| Asthma | 3,119 (15.0) | 0 | 3,119 (23) | 1,126 (13.3) | 1,993 (38.9) |
| Bronchitis | 552 (2.7) | 0 | 552 (4.1) | 115 (1.4) | 437 (8.5) |
| Diabetes | 6,687 (32.2) | 0 | 6,687 (49.2) | 2,751 (32.5) | 3,936 (76.8) |
| Hypertension/high blood pressure | 9,481 (45.6) | 0 | 9,481 (59.8) | 4,712 (55.7) | 4,769 (93.0) |
| Low blood pressure | 5,112 (24.6) | 0 | 5,112 (37.6) | 1,616 (19.1) | 3,496 (68.2) |
| Overweight or obesity | 5,438 (26.1) | 0 | 5,438 (40.0) | 1,817 (21.5) | 3,621 (70.6) |
| Rheumatoid arthritis | 172 (0.8) | 0 | 172 (1.3) | 57 (0.7) | 115 (2.2) |

Notes: <sup>a</sup> Proportions for area-level deprivation were calculated from available data

The most prevalent conditions were hypertension (46%), followed by diabetes (32%), overweight or obesity (26%), low blood pressure (25%) and asthma (15%); bronchitis (3%) and rheumatoid arthritis (1%) were not common (Table 1). Information on the sociodemographic and clinical factors stratified by ethnicity are presented in Table 2.

**Table 2.** Ethnic differences in the sociodemographic factors and health conditions

| % within ethnicity | White British | Black British | Black African | Black Caribbean | Other White | Asian British | Mixed ethnicity | Irish | Indian | Pakistani | Chinese | Bangladeshi | Other ethnicity | p-value |
| --- | --- | --- | --- | --- | --- | --- | --- | --- | --- | --- | --- | --- | --- | --- |
| Women (Gender) | 38.1 | 38.7 | 42.7 | 40.5 | 38.5 | 39.4 | 43.8 | 29.1 | 39.5 | 43.5 | 60.5 | 40.8 | 39.1 | <.001 |
| Age: *Mdn (IQR)* | 39.4 (28.6 - 50.5) | 32.7 (24.0 - 42.9) | 34.9 (26.3 - 43.9) | 42.8 (31.9 - 50.5) | 35.3 (28.0 - 44.2) | 36.1 (26.7 - 45.3) | 29.8 (22.0 - 40.6) | 43.0 (32.5 - 52.9) | 41.3 (32.1 - 51.3) | 32.6 (24.9 - 43.1) | 32.1 (26.3 - 41.7) | 31.0 (25.3 - 39.2) | 35.0 (27.0 - 45.6) |  |
| 13-17 years | 5.0 | 6.7 | 3.8 | 3.3 | 2.7 | 5.1 | 12.8 | 3.9 | 3.4 | 7.25 | 4.5 | 4.8 | 3.3 | <.001 |
| 18-34 years | 34.0 | 48.4 | 46.5 | 27.0 | 46.5 | 42.1 | 48.6 | 28.5 | 29.7 | 48.8 | 55.2 | 60.0 | 46.7 |  |
| 35-49 years | 34.7 | 35.6 | 37.1 | 43.0 | 35.1 | 35.7 | 28.6 | 37.5 | 37.9 | 28.0 | 28.4 | 28.8 | 32.9 |  |
| 50-65 years | 26.3 | 9.3 | 12.7 | 26.7 | 15.7 | 17.1 | 10.0 | 30.0 | 29.1 | 15.9 | 11.9 | 6.4 | 17.1 |  |
| Area-level deprivation |  |  |  |  |  |  |  |  |  |  |  |  |  | <.001 |
| Least deprived (7-10 deciles) | 12.9 | 3.8 | 2.6 | 3.6 | 8.6 | 8.2 | 8.8 | 6.5 | 6.4 | 4.6 | 9.2 | 7.8 | 5.4 |  |
| Middle deprivation (5-6 deciles) | 18.7 | 15.0 | 11.0 | 15.5 | 17.9 | 14.8 | 15.5 | 15.7 | 22.0 | 25.8 | 17.7 | 14.8 | 15.2 |  |
| Second most deprived (3-4 deciles) | 34.7 | 40.7 | 38.8 | 38.8 | 37.1 | 41.3 | 40.3 | 40.82 | 45.5 | 43.9 | 34.5 | 30.4 | 40.4 |  |
| Most deprived (1-2 deciles) | 33.7 | 40.5 | 47.6 | 42.1 | 36.4 | 35.7 | 35.4 | 37.1 | 26.1 | 25.8 | 38.7 | 47.0 | 39.0 |  |
| Observation period (years): *M (SD)* | 7.78 (4.42) | 8.44 (4.44) | 8.56 (4.33) | 9.70 (4.29) | 7.52 (4.31) | 8.26 (4.28) | 7.82 (4.59) | 8.37 (4.28) | 8.46 (4.40) | 7.32 (4.49) | 8.70 (4.04) | 8.04 (4.12) | 6.81 (3.82) | <.001 |
| *Physical health comorbidities* |  |  |  |  |  |  |  |  |  |  |  |  |  |  |
| Asthma | 16.2 | 19.9 | 11.2 | 17.9 | 10.3 | 12.0 | 18.6 | 17.4 | 15.3 | 12.6 | 6.7 | 10.4 | 8.1 | <.001 |
| Bronchitis | 4.0 | 2.0 | 1.5 | 5.9 | 2.0 | 2.0 | 2.0 | 4.2 | 1.8 | 2.4 | 1.5 | 0.8 | 0.7 | <.001 |
| Diabetes | 27.7 | 38.9 | 35.9 | 46.5 | 23.8 | 35.9 | 33.0 | 28.5 | 39.8 | 30.4 | 22.4 | 34.4 | 20.4 | <.001 |
| Hypertension | 41.8 | 53.7 | 53.0 | 60.3 | 36.4 | 40.9 | 44.0 | 41.4 | 49.2 | 35.3 | 35.1 | 34.4 | 30.3 | <.001 |
| Low blood pressure | 22.2 | 31.5 | 26.9 | 28.7 | 21.4 | 25.0 | 25.5 | 22.8 | 20.5 | 21.7 | 24.6 | 17.6 | 16.5 | <.001 |
| Overweight or obesity | 22.2 | 33.6 | 30.9 | 32.6 | 22.0 | 25.4 | 25.1 | 23.1 | 22.0 | 24.6 | 20.9 | 28.8 | 19.8 | <.001 |
| Rheumatoid arthritis | 1.1 | 0.7 | 0.6 | 1.2 | 0.6 | 0.5 | 1.0 | 0.9 | 0.6 | 1.0 | 0.0 | 00.0 | 0.6 | .165 |

### Ethnicity and odds for multimorbidity

The results from the logistic regression analyses (Table 3) showed that adjusting for age, gender, area-level deprivation and observation period, compared to White British people, Black African [aOR=1.41, 95%CI (1.23 - 1.56)], Black Caribbean [aOR=1.79, 95%CI (1.58 – 2.03)], and Black British people [aOR=1.64, 95%CI (1.49 – 1.81)] were more likely to have multimorbidity (psychosis plus one physical health condition). Reduced odds for multimorbidity was observed among people of Chinese [aOR=0.61, 95%CI (0.43 – 0.88)] and Other ethnicities [aOR=0.67, 95%CI (0.59 – 0.76)]. The magnitude of these differences was even higher when comparing the odds for severe multimorbidity, having 3 or more physical health conditions (Table 3). No significant differences in odds for multimorbidity were observed between White British and people of South Asian background or Asian British, Irish, Other White background, and mixed race. However, in the model regarding multimorbidity severity, people of White Other ethnicity were observed to have reduced odds for having 3 or more physical health conditions [aOR=0.77, 95%CI (0.66 – 0.91)], compared to White British people. The effects of ethnicity on multimorbidity did not differ by gender or level of deprivation as indicated by the statistically non-significant interaction terms in the logistic models [likelihood ratio (LR) test based chi-squared = 14.26(12), *p* = 0.284 for ethnicity x gender and 44.48(36), *p* = 0.157 for ethnicity x deprivation interactions respectively].

**Table 3.**
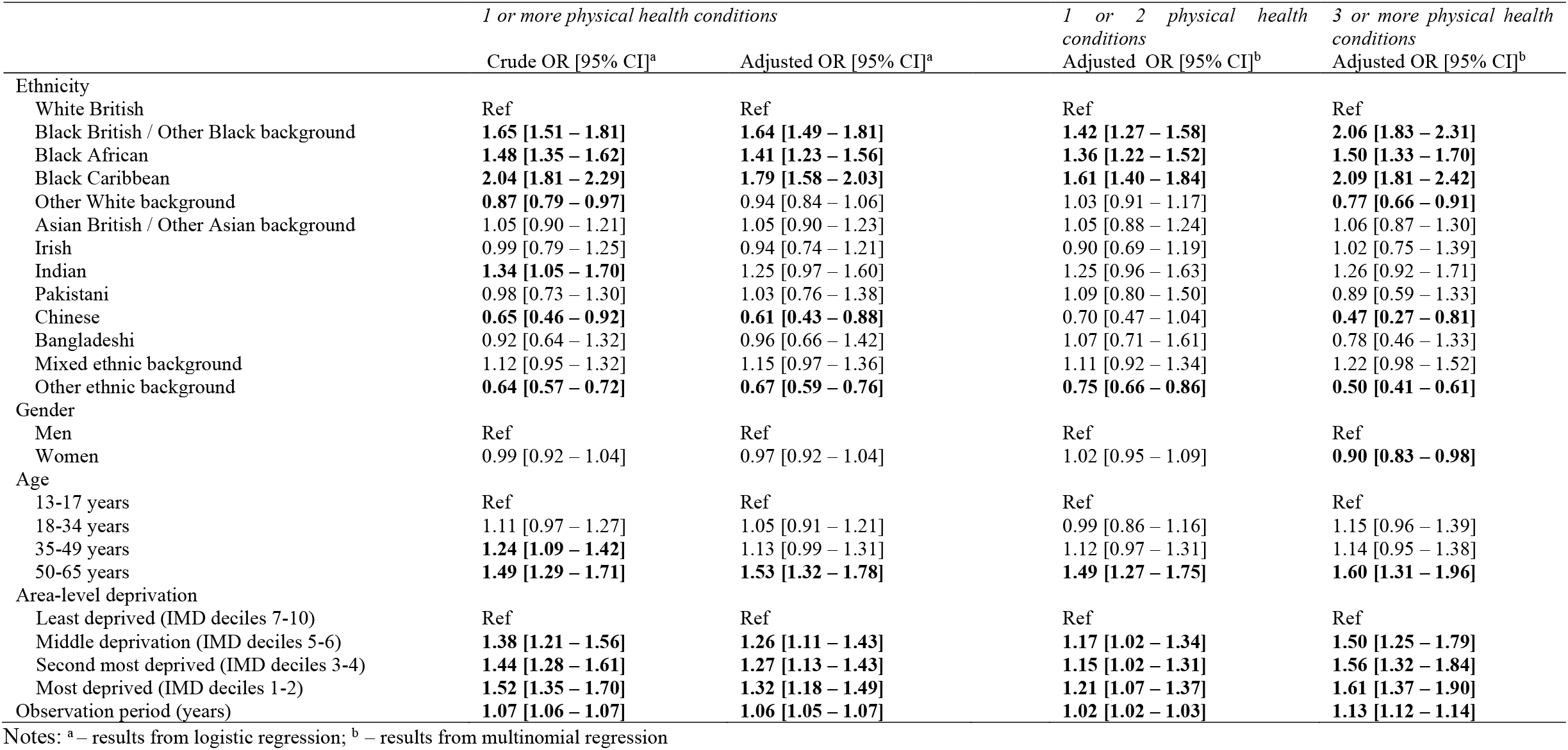
Odd ratios (OR) for having multimorbidity compared to having no physical health conditions

## DISCUSSION

We investigated ethnic inequities in risks of multimorbidity in people with psychosis, using data from the electronic health records of South London and Maudsley NHS Trust. Black African, Black Caribbean and Black British people with psychosis have around 1.5 greater odds for multimorbidity, and around twice the odds for severe multimorbidity (i.e. psychosis plus three physical health conditions). People of Chinese ethnicity and Other ethnicity had reduced odds for multimorbidity. This is the first study that investigates potential ethnic inequities in multimorbidity among a cohort of people with psychosis. These findings are partly in line with a study that observed a greater risk of the development of diabetes among minoritised ethnic people within the first year of treatment for psychosis (Gaughran et al., 2019). Also, the pattern of disparities observed is partially similar to a study of multimorbidity among a national sample of older adults (Watkinson et al., 2021). Similar findings are the greater risk observed among Black people, the reduced odds among Chinese people, and the absence of differences among people with Mixed ethnicity (Watkinson et al., 2021). However, in our study, we did not observe an increased odds for multimorbidity among people of South Asian, Asian British, or Other ethnicities. The age difference of the samples and the fact that we only investigated six health conditions, while the study of multimorbidity among older adults investigated 14 (Watkinson et al., 2021), may be the reasons for differences in findings.

The observed ethnic inequities could probably be driven by inequities in the social determinants of health beyond the included measure of area deprivation, including social marginalisation, racial discrimination, and, for migrants, the stresses related to migration and acculturation (Allen et al., 2014; Bhui et al., 2021; Cabinet Office, 2018; Hatch & Dohrenwend, 2007; Jongsma et al., 2020; Marmot et al., 2020; James Y. Nazroo et al., 2020). For instance, experiences of racism and racial discrimination have been related to worse mental and physical health, including the onset of psychosis, diabetes, respiratory illness, and hypertension (Jongsma et al., 2020; Karlsen et al., 2005; Karlsen & Nazroo, 2002; Paradies et al., 2015; Pearce et al., 2019; Williams et al., 2019). Health-related behaviours could mediate the relationships between social adversity and the onset of multimorbidity (Bhui et al., 2021; Williams et al., 2019). The findings may also reflect the shared aetiology of mental and physical health conditions. One study has reported an association between the number of psychotic symptoms and number of physical health conditions (Moreno et al., 2013). The role of social stress, allostatic load, bodily inflammation and coagulation have been related to the onset of physical health disorders, as well as psychosis; these mechanisms may explain multimorbidity with psychosis (Beckie, 2012; Heurich et al., 2021; Howes & McCutcheon, 2017; Lund et al., 2018).

### Strengths and limitations

This study used data from one of the UK’s largest secondary mental health facilities, serving an ethnically diverse community. The cohort of the study is representative of South London and Maudsley NHS Trust catchment, and the findings should be generalisable to other ethnically dense urban areas in the United Kingdom. Indeed, most ethnic minorities live in urban areas. Ethnicity was self-defined, however we could not include other relevant factors that can be related to inequities in health, namely migration and potential related barriers in the use of health services. Also, we could not adjust for individual measures socio-economic status (e.g., income, level of education and experiences of racism and discrimination) which have been associated with health inequities (Karlsen & Nazroo, 2002). There are limitations in using Natural Language Processing (NLP) algorithms to assess the prevalence of physical health conditions, however the use of NLP enabled us to investigate ethnic inequities in a larger sample of people than would be possible if restricting to people whose data could be linked to other administrative datasets that contain data on physical morbidity, such as the Lambeth DataNet (Woodhead et al., 2014). There is also the risk of underestimation of the prevalence of some physical health conditions, as they would be managed in primary care and not recorded in SLaM (Gaughran et al., 2019). We did not account for potential ethnic differences in psychotic illness severity and healthcare service use (Chui et al., 2021; Morris et al., 2020), which may affect the availability of information. Moreover, only the conditions relevant to the aetiology and management of psychosis may be reported in SLaM electronic records, so we may have less information on other conditions. We did not adjust for the type of pharmacological treatment for psychosis, which could be related to the presence of the included physical conditions (Baxter et al., 2016; Dubath et al., 2021). Finally, we could not investigate the timing of the onset of the different health conditions. Future cohort studies are needed to verify and expand on our findings (e.g., Das-Munshi et al., 2016).

## CONCLUSIONS

This study shows that Black people with psychosis are at higher risk of multimorbidity. Although this study is not focused on aetiology, the findings are consistent with adversity-related onset of disease, where marginalised ethnic groups face higher rates of psychosis and other health conditions when living with this severe mental illness (Dubath et al., 2021; Jongsma et al., 2021; Watkinson et al., 2021). The observed ethnic inequities in health show the need for integrated care and multi-disease prevention among minoritised ethnic groups.

## Data Availability

The data that support the findings of this study are available on request from the corresponding author. The data are not publicly available due to the Information Governance framework and Research Ethics Committee approval in place concerning CRIS data use.

## STATEMENTS

### Ethical Standards

CRIS dataset received approval from the Oxford C Research Ethics Committee (18/SC/0372). All projects using the CRIS dataset are submitted for approval to an oversight committee led by service-users (this project reference: 20-075). Service-users consent for publication of studies using CRIS data is not required.

### Authors’ contributions

KB developed the original idea in relation to new work on syndemics, and developed this with DFdF. KB, DFdF, MK, JN and RDH were involved in the reviewing the methodology and analytic plans. MP and HS curated the data. DFdF undertook all data analyses and wrote the first draft. All authors provided critical insight, reviewed, and suggested edits and agreed to the final version.

## Footnotes

1 We use the term inequity in line with the World Health Organisation definition of health equity as “the absence of unfair, avoidable or remediable differences among groups of people, whether those groups are defined socially, economically, demographically, or geographically or by other dimensions of inequality (e.g., sex, gender, ethnicity, disability, or sexual orientation).” (World Health Organization, 2021).

## REFERENCES

Allen, J., Balfour, R., Bell, R., & Marmot, M. (2014). Social determinants of mental health. International Review of Psychiatry, 26(4), 392–407. https://doi.org/10.3109/09540261.2014.928270

Baxter, A. J., Harris, M. G., Khatib, Y., Brugha, T. S., Bien, H., & Bhui, K. (2016). Reducing excess mortality due to chronic disease in people with severe mental illness: Meta-review of health interventions. British Journal of Psychiatry, 208(4), 322–329. https://doi.org/10.1192/bjp.bp.115.163170

Beckie, T. M. (2012). A Systematic Review of Allostatic Load, Health, and Health Disparities. Biological Research for Nursing, 14(4), 311–346. https://doi.org/10.1177/1099800412455688

Benchimol, E. I., Smeeth, L., Guttmann, A., Harron, K., Moher, D., Petersen, I., Sørensen, H. T., von Elm, E., & Langan, S. M. (2015). The REporting of studies Conducted using Observational Routinely-collected health Data (RECORD) Statement. PLOS Medicine, 12(10), e1001885. https://doi.org/10.1371/journal.pmed.1001885

Bhui, P. K., Havorsrud, K., Mooney, R., & Hosang, G. M. (2021). Is psychosis a syndemic manifestation of historical and contemporary adversity ? Findings from UKBioBank. The British Journal of Psychiatry. https://doi.org/10.1192/bjp.2021.142

Bisquera, A., Turner, E. B., Ledwaba-Chapman, L., Dunbar-Rees, R., Hafezparast, N., Gulliford, M., Durbaba, S., Soley-Bori, M., Fox-Rushby, J., Dodhia, H., Ashworth, M., & Wang, Y. (2021). Inequalities in developing multimorbidity over time: A population-based cohort study from an urban, multi-ethnic borough in the United Kingdom. The Lancet Regional Health - Europe, 12, 100247. https://doi.org/10.1016/j.lanepe.2021.100247

Cabinet Office. (2018). Race Disparity Audit. https://assets.publishing.service.gov.uk/government/uploads/system/uploads/attachment_data/file/686071/Revised_RDA_report_March_2018.pdf

Chui, Z., Gazard, B., MacCrimmon, S., Harwood, H., Downs, J., Bakolis, I., Polling, C., Rhead, R., & Hatch, S. L. (2021). Inequalities in referral pathways for young people accessing secondary mental health services in south east London. European Child & Adolescent Psychiatry, 30(7), 1113–1128. https://doi.org/10.1007/s00787-020-01603-7

CRIS NLP Service. (2021). Library of production-ready applications, v1.6. https://www.maudsleybrc.nihr.ac.uk/facilities/clinical-record-interactive-search-cris/cris-natural-language-processing/

Das-Munshi, J., Ashworth, M., Gaughran, F., Hull, S., Morgan, C., Nazroo, J., Roberts, A., Rose, D., Schofield, P., Stewart, R., Thornicroft, G., & Prince, M. J. (2016). Ethnicity and cardiovascular health inequalities in people with severe mental illnesses: protocol for the E-CHASM study. Social Psychiatry and Psychiatric Epidemiology, 51(4), 627–638. https://doi.org/10.1007/s00127-016-1185-8

Department for Communities and Local Government. (2015). The English Indices of Deprivation 2015 - Technical Report (Issue September). https://www.gov.uk/government/uploads/system/uploads/attachment_data/file/464485/English_Indices_of_Deprivation_2015_-_Technical-Report.pdf

Dubath, C., Gholam-Rezaee, M., Sjaarda, J., Levier, A., Saigi-Morgui, N., Delacrétaz, A., Glatard, A., Panczak, R., Correll, C. U., Solida, A., Plessen, K. J., von Gunten, A., Kutalik, Z., Conus, P., & Eap, C. B. (2021). Socio-economic position as a moderator of cardiometabolic outcomes in patients receiving psychotropic treatment associated with weight gain: results from a prospective 12-month inception cohort study and a large population-based cohort. Translational Psychiatry, 11(1), 360. https://doi.org/10.1038/s41398-021-01482-9

El-Sayed, A. M., Scarborough, P., & Galea, S. (2011). Ethnic inequalities in obesity among children and adults in the UK: a systematic review of the literature. Obesity Reviews, 12(5), e516–e534. https://doi.org/10.1111/j.1467-789X.2010.00829.x

Evandrou, M., Falkingham, J., Feng, Z., & Vlachantoni, A. (2016). Ethnic inequalities in limiting health and self-reported health in later life revisited. Journal of Epidemiology and Community Health, 70(7), 653–662. https://doi.org/10.1136/jech-2015-206074

Gaughran, F., Stahl, D., Stringer, D., Hopkins, D., Atakan, Z., Greenwood, K., Patel, A., Smith, S., Gardner-Sood, P., Lally, J., Heslin, M., Stubbs, B., Bonaccorso, S., Kolliakou, A., Howes, O., Taylor, D., Di Forti, M., David, A. S., Murray, R. M., & Ismail, K. (2019). Effect of lifestyle, medication and ethnicity on cardiometabolic risk in the year following the first episode of psychosis: Prospective cohort study. British Journal of Psychiatry, 215(6), 712–719. https://doi.org/10.1192/bjp.2019.159

Hatch, S. L., & Dohrenwend, B. P. (2007). Distribution of traumatic and other stressful life events by race/ethnicity, gender, SES and age: A review of the research. American Journal of Community Psychology, 40(3–4), 313–332. https://doi.org/10.1007/s10464-007-9134-z

Head, A., Fleming, K., Kypridemos, C., Schofield, P., Pearson-Stuttard, J., & O’Flaherty, M. (2021). Inequalities in incident and prevalent multimorbidity in England, 2004–19: a population-based, descriptive study. The Lancet Healthy Longevity, 2(8), e489–e497. https://doi.org/10.1016/S2666-7568(21)00146-X

Heurich, M., Föcking, M., Mongan, D., Cagney, G., & Cotter, D. R. (2021). Dysregulation of complement and coagulation pathways: emerging mechanisms in the development of psychosis. Molecular Psychiatry, November 2020, 1–14. https://doi.org/10.1038/s41380-021-01197-9

Howes, O. D., & McCutcheon, R. (2017). Inflammation and the neural diathesis-stress hypothesis of schizophrenia: a reconceptualization. Translational Psychiatry, 7(2), e1024–e1024. https://doi.org/10.1038/tp.2016.278

Jackson, R. G., Patel, R., Jayatilleke, N., Kolliakou, A., Ball, M., Gorrell, G., Roberts, A., Dobson, R. J., & Stewart, R. (2017). Natural language processing to extract symptoms of severe mental illness from clinical text: the Clinical Record Interactive Search Comprehensive Data Extraction (CRIS-CODE) project. BMJ Open, 7(1), e012012. https://doi.org/10.1136/bmjopen-2016-012012

Johnston, M. C., Crilly, M., Black, C., Prescott, G. J., & Mercer, S. W. (2019). Defining and measuring multimorbidity: A systematic review of systematic reviews. European Journal of Public Health, 29(1), 182–189. https://doi.org/10.1093/eurpub/cky098

Jongsma, H. E., Gayer-Anderson, C., Tarricone, I., Velthorst, E., van der Ven, E., Quattrone, D., di Forti, M., Menezes, P. R., Del-Ben, C. M., Arango, C., Lasalvia, A., Berardi, D., La Cascia, C., Bobes, J., Bernardo, M., Sanjuán, J., Santos, J. L., Arrojo, M., de Haan, L., … Kirkbride, J. B. (2020). Social disadvantage, linguistic distance, ethnic minority status and first-episode psychosis: results from the EU-GEI case–control study. Psychological Medicine, 1–13. https://doi.org/10.1017/S003329172000029X

Jongsma, H. E., Karlsen, S., Kirkbride, J. B., & Jones, P. B. (2021). Understanding the excess psychosis risk in ethnic minorities: the impact of structure and identity. Social Psychiatry and Psychiatric Epidemiology, 0123456789. https://doi.org/10.1007/s00127-021-02042-8

Karlsen, S., & Nazroo, J. Y. (2002). Relation between racial discrimination, social class, and health among ethnic minority groups. American Journal of Public Health, 92(4), 624–631. http://www.pubmedcentral.nih.gov/articlerender.fcgi?artid=1447128&tool=pmcentrez&rendertype=abstract

Karlsen, S., Nazroo, J. Y., McKenzie, K., Bhui, K., & Weich, S. (2005). Racism, psychosis and common mental disorder among ethnic minority groups in England. Psychological Medicine, 35(12), 1795–1803. https://doi.org/10.1017/S0033291705005830

Larsen, F. B., Pedersen, M. H., Friis, K., Glümer, C., & Lasgaard, M. (2017). A latent class analysis of multimorbidity and the relationship to socio-demographic factors and health-related quality of life. A national population-based study of 162,283 Danish adults. PLOS ONE, 12(1), e0169426. https://doi.org/10.1371/journal.pone.0169426

Lund, C., Brooke-Sumner, C., Baingana, F., Baron, E. C., Breuer, E., Chandra, P., Haushofer, J., Herrman, H., Jordans, M., Kieling, C., Medina-Mora, M. E., Morgan, E., Omigbodun, O., Tol, W., Patel, V., & Saxena, S. (2018). Social determinants of mental disorders and the Sustainable Development Goals: A systematic review of reviews. The Lancet Psychiatry, 5(4), 357–369. https://doi.org/10.1016/S2215-0366(18)30060-9

Marmot, M., Allen, J., Boyce, T., Goldblatt, P., & Morrison, J. (2020). Health equity in England: The Marmot Review 10 years on. <http://www.instituteofhealthequity.org/resources-reports/marmot-review-10-years-on>

Moreno, C., Nuevo, R., Chatterji, S., Verdes, E., Arango, C., & Ayuso-Mateos, J. L. (2013). Psychotic symptoms are associated with physical health problems independently of a mental disorder diagnosis: Results from the WHO World Health Survey. World Psychiatry, 12(3), 251–257. https://doi.org/10.1002/wps.20070

Morris, R. M., Sellwood, W., Edge, D., Colling, C., Stewart, R., Cupitt, C., & Das-Munshi, J. (2020). Ethnicity and impact on the receipt of cognitive–behavioural therapy in people with psychosis or bipolar disorder: an English cohort study. BMJ Open, 10(12), e034913. https://doi.org/10.1136/bmjopen-2019-034913

Nazroo, J. Y., Falaschetti, E., Pierce, M., & Primatesta, P. (2009). Ethnic inequalities in access to and outcomes of healthcare: analysis of the Health Survey for England. Journal of Epidemiology and Community Health, 63(12), 1022–1027. https://doi.org/10.1136/jech.2009.089409

Nazroo, James Y. (1998). Genetic, cultural or socio-economic vulnerability? Explaining ethnic inequalities in health. Sociology of Health & Illness, 20(5), 710–730. https://doi.org/10.1111/1467-9566.00126

Nazroo, James Y., Bhui, K. S., & Rhodes, J. (2020). Where next for understanding race/ethnic inequalities in severe mental illness? Structural, interpersonal and institutional racism. Sociology of Health & Illness, 42(2), 262–276. https://doi.org/10.1111/1467-9566.13001

Nazroo, James Y., & Williams, D. R. (2006). The social determination of ethnic/racial inequalities in health. In M. Marmot & R. G. Wilkinson (Eds.), Social Determinants of Health (pp. 238–266). Oxford University Press. https://doi.org/10.1093/acprof:oso/9780198565895.003.12

NICE (National Institute for Health and Care Excellence). (2016). Multimorbidity: clinical assessment and management (Issue September 2016). http://www.nice.org.uk/guidance/ng56

Paradies, Y., Ben, J., Denson, N., Elias, A., Priest, N., Pieterse, A., Gupta, A., Kelaher, M., & Gee, G. (2015). Racism as a determinant of health: A systematic review and meta-analysis. PLOS ONE, 10(9), 1–48. https://doi.org/10.1371/journal.pone.0138511

Pearce, J., Rafiq, S., Simpson, J., & Varese, F. (2019). Perceived discrimination and psychosis: a systematic review of the literature. Social Psychiatry and Psychiatric Epidemiology, 54(9), 1023–1044. https://doi.org/10.1007/s00127-019-01729-3

Perera, G., Broadbent, M., Callard, F., Chang, C. K., Downs, J., Dutta, R., Fernandes, A., Hayes, R. D., Henderson, M., Jackson, R., Jewell, A., Kadra, G., Little, R., Pritchard, M., Shetty, H., Tulloch, A., & Stewart, R. (2016). Cohort profile of the South London and Maudsley NHS Foundation Trust Biomedical Research Centre (SLaM BRC) Case Register: current status and recent enhancement of an Electronic Mental Health Record-derived data resource. BMJ Open, 6(3), 1–22. https://doi.org/10.1136/bmjopen-2015-008721

Rodrigues, M., Wiener, J. C., Stranges, S., Ryan, B. L., & Anderson, K. K. (2021). The risk of physical multimorbidity in people with psychotic disorders: A systematic review and meta-analysis. Journal of Psychosomatic Research, 140, 110315. https://doi.org/10.1016/j.jpsychores.2020.110315

Schofield, P., Saka, O., & Ashworth, M. (2011). Ethnic differences in blood pressure monitoring and control in south east London. British Journal of General Practice, 61(585), e190–e196. https://doi.org/10.3399/bjgp11X567126

Sproston, K., & Mindell, J. (2006). The health of minority ethnic groups. http://www.ic.nhs.uk/webfiles/publications/healthsurvey2004ethnicfull/HealthSurveyforEnglandVol1_210406_PDF.pdf

StataCorp. (2017). Stata Statistical Software: Release 15. StataCorp LLC.

Stewart, R., Soremekun, M., Perera, G., Broadbent, M., Callard, F., Denis, M., Hotopf, M., Thornicroft, G., & Lovestone, S. (2009). The South London and Maudsley NHS Foundation Trust Biomedical Research Centre (SLAM BRC) case register: development and descriptive data. BMC Psychiatry, 9(1), 51. https://doi.org/10.1186/1471-244X-9-51

Stubbs, B., Koyanagi, A., Veronese, N., Vancampfort, D., Solmi, M., Gaughran, F., Carvalho, A. F., Lally, J., Mitchell, A. J., Mugisha, J., & Correll, C. U. (2016). Physical multimorbidity and psychosis: Comprehensive cross sectional analysis including 242,952 people across 48 low-and middle-income countries. BMC Medicine, 14(1), 1–12. https://doi.org/10.1186/s12916-016-0734-z

Verest, W. J. G. M., Galenkamp, H., Spek, B., Snijder, M. B., Stronks, K., & van Valkengoed, I. G. M. (2019). Do ethnic inequalities in multimorbidity reflect ethnic differences in socioeconomic status? The HELIUS study. European Journal of Public Health, 29(4), 687–693. https://doi.org/10.1093/eurpub/ckz012

Watkinson, R. E., Sutton, M., & Turner, A. J. (2021). Ethnic inequalities in health-related quality of life among older adults in England: secondary analysis of a national cross-sectional survey. The Lancet Public Health, 6(3), e145–e154. https://doi.org/10.1016/S2468-2667(20)30287-5

Williams, D. R., Lawrence, J. A., Davis, B. A., & Vu, C. (2019). Understanding how discrimination can affect health. Health Services Research, 54(S2), 1374–1388. https://doi.org/10.1111/1475-6773.13222

Woodhead, C., Ashworth, M., Schofield, P., & Henderson, M. (2014). Patterns of physical co-/multi-morbidity among patients with serious mental illness: A London borough-based cross-sectional study. BMC Family Practice, 15(1), 1–9. https://doi.org/10.1186/1471-2296-15-117

World Health Organization. (2021). Health Equity. https://www.who.int/health-topics/health-equity

